# COVID-19 Confirmed Cases and Fatalities in 883 U.S. Counties with a Population of 50,000 or More: Predictions Based on Social, Economic, Demographic Factors and Shutdown Days

**DOI:** 10.1101/2020.06.25.20139956

**Authors:** Leon S. Robertson

## Abstract

The spread of the COVID-19 virus is highly variable among U.S. counties. Seventeen factors known or thought to be related to spread of the COVID-19 virus were studied by Poisson regression analysis of confirmed cases and deaths in 883 U.S. counties with a population of 50,000 or more as of May 31, 2020. With little exception, each factor was predictive of incidence and mortality. The regression equation can be used to identify priority locations for preventive efforts and preparation for medical care caseloads when prevention is unsuccessful. Based on the correlation of cases and deaths to days since stay-at-home orders were issued, the orders reduced the cases about 48 percent and deaths about 50 percent. Focusing preventive efforts on the more vulnerable counties may be more effective and less economically damaging than statewide shutdowns.

## INTRODUCTION

Various mathematical models using different methods initially produced quite different predictions of COVID-19 cases and deaths in the U.S. because of the variance in assumptions about how the virus and people would behave. As more data became available the projections converged somewhat but still varied substantially (1). The early predictions did serve the purpose of motivating U.S. states to adopt policies to slow the spread of the virus. In the U.S. the timing of stay-at-home (shutdown) orders varied among the states. The government and business operations and other gatherings prohibited during the shutdown also varied somewhat among the states but all left plenty of leeway for the virus to continue to spread. Several state governors did not issue such orders and many announced partial or complete termination of the orders in late April and early May, 2020. In Wisconsin and Oregon the order was voided by judges. Some state and local governments issued standards for physical distancing in businesses and other organization as well as wearing masks in public places after the shutdown but enforcement will be problematic as was the stay-at-home orders. If local medical care facilities are to be prepared for an influx of cases, it is important to be able to predict the rise in numbers in local areas as early as possible. Large swaths of counties in the U.S. have no hospital beds, and if they have them, no intensive care units. More than half of U.S. counties have no intensive care beds (2).

Human interaction is obviously a major factor in the spread of a human transported infectious agent. COVID-19 virus is spread by breathing, coughing, sneezing, talking and singing as well as touching surfaces by those infected. In response to that information, behavior in the U.S. varied from substantial recommended risk avoidance behavior (e.g., reduced travel, reduced physical proximity to other people, frequent hand washing, wearing face masks) to mockery of those who did so and protests against requirements to do so (3). Photographs and videos of street protesters against shutdowns showed many people in close proximity to one another with no face masks (4). The virus was well established in many communities before testing, tracking those exposed and quarantine was initiated.

The authors of one study claimed that the warnings and shutdowns prevented 60 million COVID-19 cases in the U.S. based on models of exponential growth in time (5). Such models applied to U.S. counties found wide variation in growth rates. Some were exponential but many others were more linear (6). A comparison of counties in Iowa with no shutdown and Illinois before and during the Illinois shutdown indicated 30 percent excess cases in Iowa counties through April 20, 2020 (7).Travel data based on tracing cell phone movements indicate that travel decreased substantially prior to the adoption of stay-at-home orders in many metropolitan areas in the U.S. but increased in time later (8) suggesting that risk avoidance began before the orders and deteriorated thereafter.

Governments and other organizations in the U.S. collect data on health, education and other characteristics of the populations, housing, businesses, religious institutions and travel specific to U.S. counties that are thought to increase or decrease the risk of human transmission of pathogens or the severity of the illnesses they cause. The purpose of this paper is to report an analysis of the extent to which available data are predictive of the counts of COVID-19 confirmed cases and deaths in U.S. counties with populations of 50,000 or more as of May 31, 2020. The study indicates that the estimate of 60 million cases prevented by the shutdowns in the U.S. is extraordinarily implausible.

## METHODS

Poisson regression models of cases and deaths in U.S. counties with 50,000 or more population were used to assess the predictive value of each factor corrected for population size and the predictive value of the other factors. The form of the regression equation is:

Number of cases (or separately, deaths) =

b1 (days from the shutdown order, if any, until May 31, 2020) +

b2 (log(population density, estimated 2019 residents per square kilometer) +

b3 (average number of persons per household) +

b4 (log(average employees per business enterprise)) +

b5 (log(average religious adherents per congregation)) +

b6 (log(average number of social acquaintances reported per person)) +

b7 (percent decline in 7-day moving average of travel per day before the shutdown)

b8 (percent of the population that is obese) +

b9 (percent of the population with diabetes) +

b10 (Medicare cardiovascular hospitalization discharges 2015-2017) +

b11 (percent of the population 65 years or older) +

b12 (percent of adults who finished high school) +

b13 (log(median family income before the pandemic)) +

b14 (income inequality before the pandemic) +

b15 (percent unemployed before the pandemic) +

b16 (Percent African American) +

b17 (Percent Hispanic)

Log(population) was included as an offset variable to correct for differences in population size among the counties. The logarithmic transformations on selected variables were used because the frequency distributions of those variables were skewed. The study was limited to counties with 50,000 or more population to avoid random variation in small numbers. To estimate the number of cases and deaths likely prevented by shutdowns, the regression equation was used to predict the number of cases and deaths in each county expected when shutdown days was set to zero. The numbers for each county were added and the totals compared to the actual number of aggregated cases and deaths.

Daily numbers of accumulated confirmed cases and deaths in each county through May 31, 2020 were downloaded from usafacts.org (https://usafacts.org/visualizations/coronavirus-covid-19-spread-map/). Dates of shutdown orders were obtained from Littler.com (https://www.littler.com/publication-press/publication/stay-top-stay-home-list-statewide). Population density was obtained from the U.S. Census Bureau (https://www.census.gov/quickfacts/fact/note/US/LND110210). Estimated 2019 population, percent unemployed and median household income prior to the pandemic for each county was downloaded from the U.S. Department of Agriculture website (https://www.ers.usda.gov/data-products/county-level-data-sets/download-data/) based on estimates from the U.S. Census Bureau and Bureau of Labor statistics. Seven day moving averages of estimated daily vehicle travel in each county from March 7 through May 31 were calculated using data provided by Streetlight Data, Inc. that relies on tracking smart phone movement. Persons per household, social acquaintances, high school graduates, economic inequality, percent 65 years or older, percent with diabetes and percent obese were downloaded from files accumulated from various sources by the Robert Wood Johnson Foundation (https://www.countyhealthrankings.org/explore-health-rankings/rankings-data-documentation). Medicare hospital discharges for cardiovascular diseases were obtained from CDC Wonder (https://nccd.cdc.gov/DHDSPAtlas/Reports.aspx). Numbers of religious adherents and congregations were obtained from therda.com (http://www.thearda.com/Archive/Files/Downloads/RCMSCY10_DL.asp). Numbers of businesses and employees were downloaded from the Bureau of Labor Statistics (https://data.bls.gov/cew/apps/table_maker/v4/table_maker.htm#type=1&year=2019&qtr=3&own=5&ind=10&supp=0). Percent African American and Hispanic were obtained from randaalolson.com (http://www.randalolson.com/2014/04/29/u-s-racial-diversity-by-county/).

## RESULTS

Data on all variables were available for 883 U.S. counties. The total population of these counties (263,489,065) was about 80 percent of the estimated 2019 U.S. population. As of May 31, 2020 confirmed COVID-19 cases in the studied counties (1,515,780) were about 85 percent of cases of the all U.S. total. Deaths attributed to COVID-19 in the studied counties as of that date were 91,649, about 90 percent of the total for all counties.

An example of the variation in growth of COVID-19 cases among counties is illustrated in Figure 1. Of the five Alabama counties displayed, only Montgomery County shows a pronounced exponential growth. Allowing for a two-week lag in time from exposure to manifestation of symptoms, the April shutdown, if effective, would have slowed the growth of COVID-19 from mid-April to mid-May. It appears to have done so in Jefferson (Birmingham) and Montgomery counties but not in Mobile. Growth in the two counties with smaller less dense populations proceeded very slowly compared to the more populous counties.

**Figure 1.**
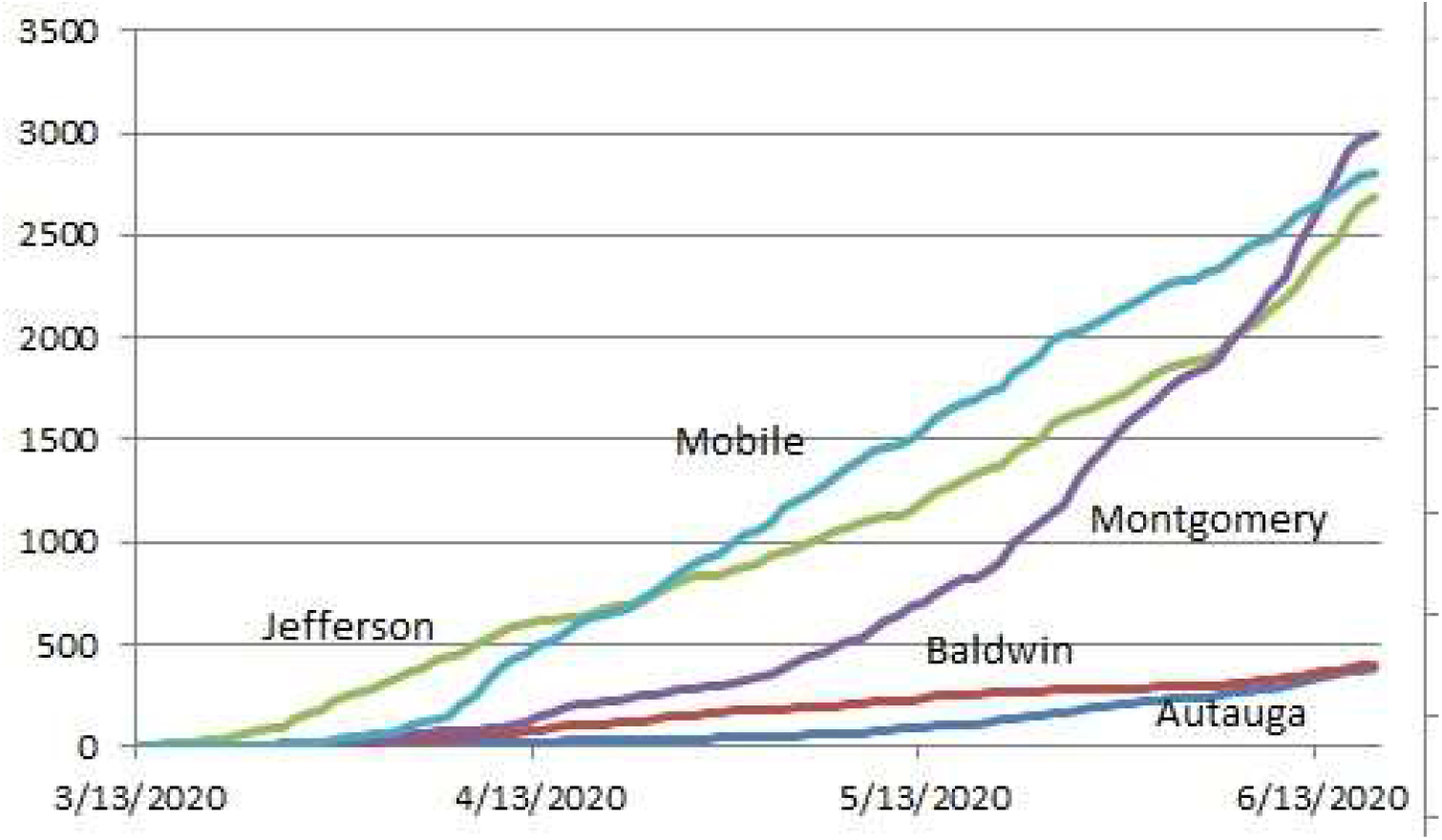
Accumulation of COVID-19 Confirmed Cases in Selected Alabama Counties.

**Figure 1.**
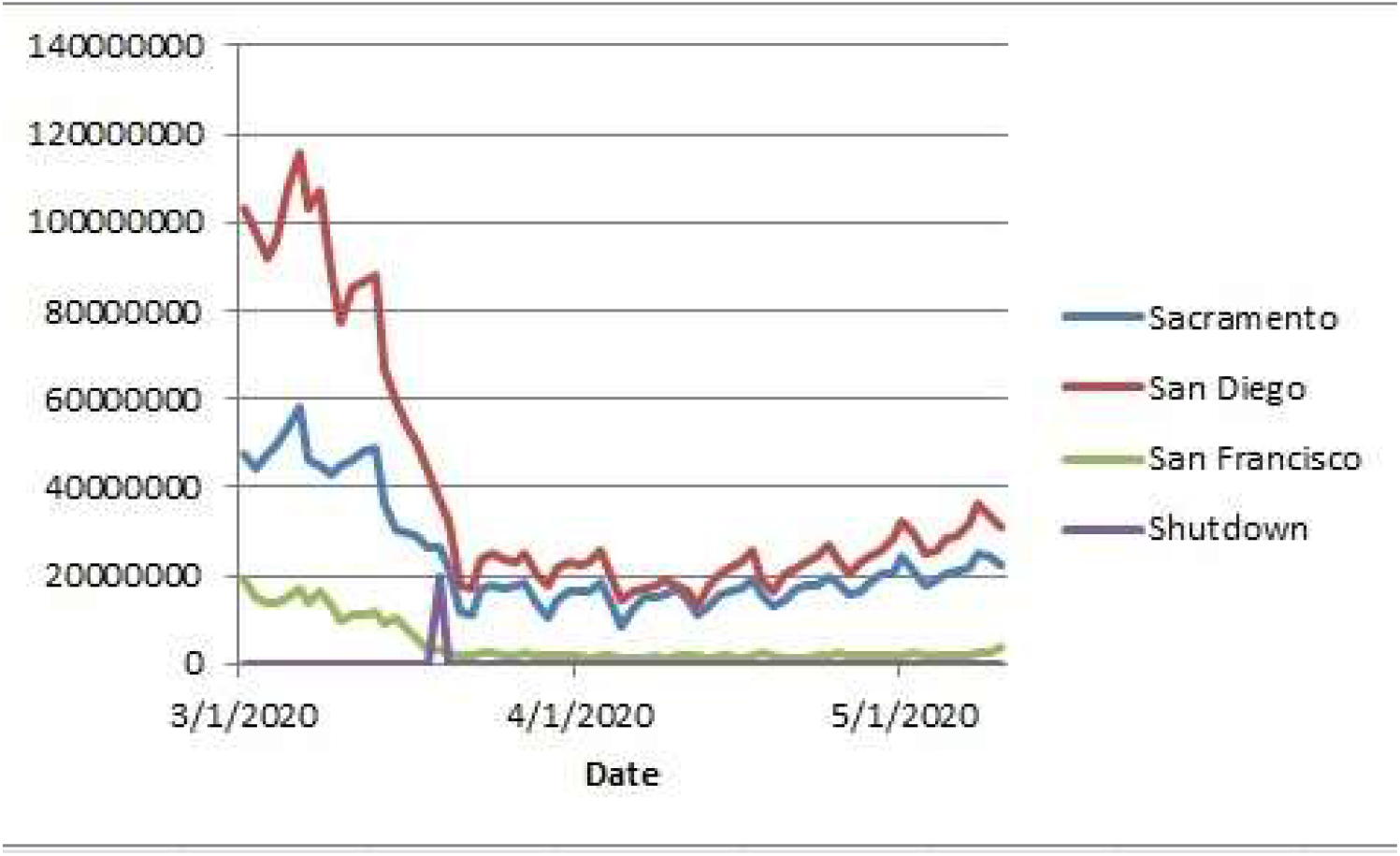
Estimated Daily Vehicle Miles Traveled in Selected California Counties.

The parameters and 95% confidence intervals of the regression estimates are shown in Table 1 separately for confirmed cases and deaths attributed to COVID-19. Most of the variables are predictive of cases and deaths with remarkably narrow confidence intervals. Counties with greater population density, crowding in housing, workplaces and religious congregations as well as self-reported social contacts had more cases and deaths. Greater numbers of cases and deaths occurred in counties where greater proportions of populations with known higher risk conditions (diabetes, heart disease and obesity) and where more elderly people live. If a lower percent of the population finished high school, cases and deaths were higher. Cases and deaths were associated with higher median incomes and higher pre-pandemic unemployment. Income inequality was associated with cases but not deaths. Cases and deaths occurred more frequently in counties with a larger proportion of African Americans in the population but were less frequent in counties with a larger proportion of Hispanics. A cautionary note: the differences among the coefficients on particular variables do not indicate greater or less importance. The variables are measured on different scales and each coefficient refers only to covariation with increments of the scale it modifies.

**Table 1.**
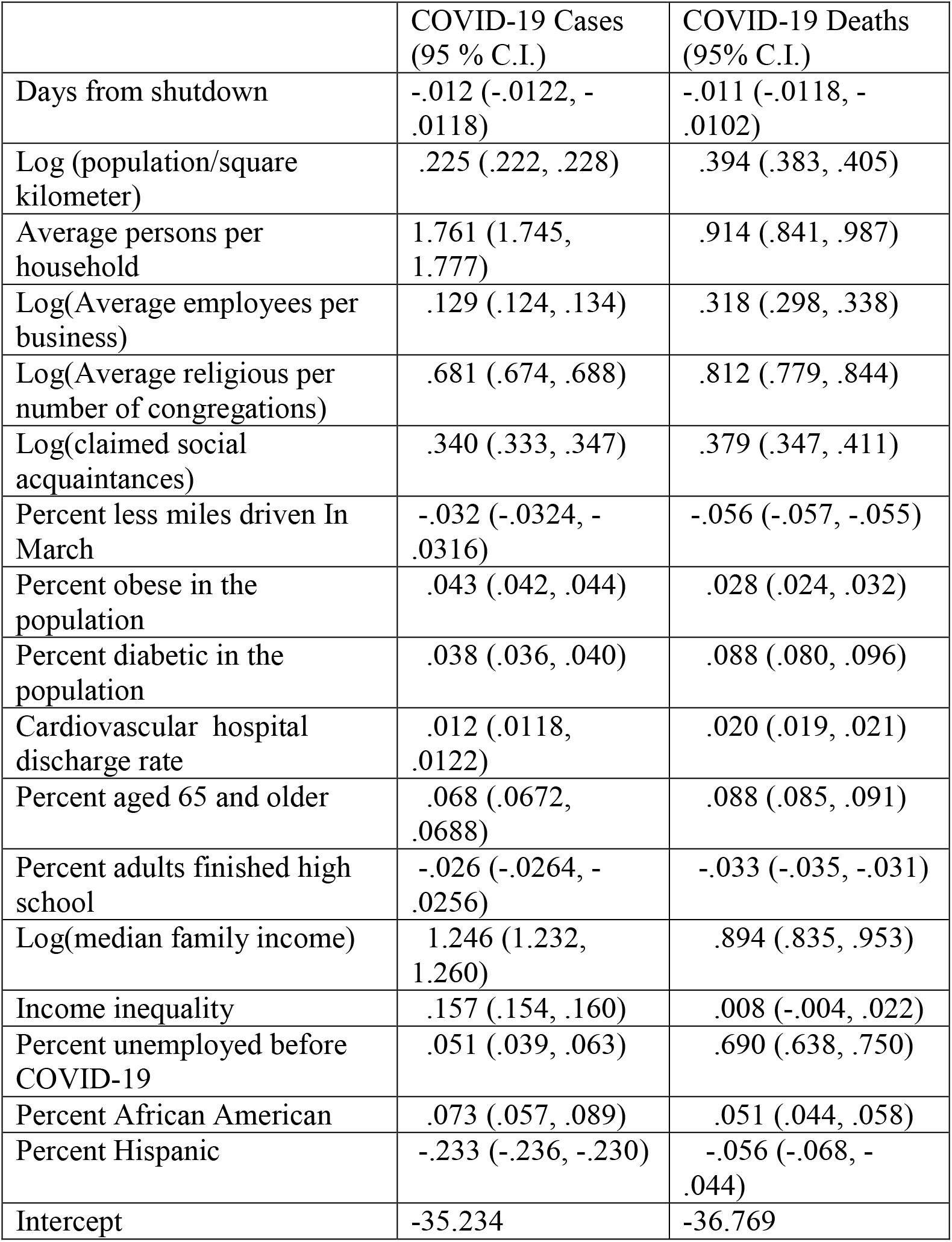
Poisson Regression Coefficients of Factors Thought to Contribute to Incidence and Severity of COVID-19 Infections, 883 U.S. Counties as of May 31, 2020

The travel variable showed a remarkable pattern. Motor vehicle travel decreased an average 68 percent in the studied counties from the first week in March through March 31, 2020. The coefficient on change in traffic is negative indicating more cases among counties where travel declined the most. That may have occurred because the increase in subsequent travel during the shutdowns was greater in counties where it declined the most. For example, Figure 1 shows the estimated miles driven in three California cities. The post-shutdown increases were proportionately greater in San Diego and Sacramento counties than in San Francisco which had a less steep decline before the shutdown.

With the shutdown coefficient set to zero, the regression model predicted 2,944,279 cases and 183,785 deaths in the studied counties through May 31, 2020. Compared to the actual 1,515,780 cases and 91,649 deaths, the results suggest that the shutdowns reduced confirmed cases by about 48 percent and deaths by about 50 percent.

## Discussion

These data further illustrate that the spread of COVID-19 is very different in local areas and is substantially predictable by numerous factors – crowding in homes, workplaces, religious gatherings -- and local economic and demographic conditions. Most of the correlations are as expected but the lower numbers of cases in counties with a larger percentage of Hispanics in the population is contrary to claims that Hispanics work disproportionately in industries where the virus spread rapidly. Since crowding in workplaces is controlled in this analysis, the coefficient on percent Hispanic is in addition to the workplace crowding variable. More detailed examination of tests and exposure tracing will be needed to explain the correlation. Similarly, the finding that the cases and deaths are more frequent in counties with higher median incomes may seem counter to the evidence that people with lower incomes are more likely to be exposed to the virus. Again the correlation prevails after controlling for other factors such as unemployment. Recall that one of the early hotspots was Westchester County New York, one of the wealthiest counties in the U.S. It is likely that air travel and business meetings increased risk among more affluent people.

The shutdowns probably prevented about 1.4 million cases and 92,000 deaths. The study that claimed the warnings and shutdowns prevented 60 million COVID-19 cases in the U.S. appears to be grossly inflated by a factor of 40. Given that travel decreased by about 68 percent when there were only a few thousand cases in the country and few shutdowns in March, 2020, it is unlikely that the numbers of cases would have increased into the tens of millions without massive voluntary risk avoidance behavior. Nevertheless, without the shutdowns, apparently the COVID-19 virus would have killed about twice as many people as had died through the end of May and caused more than enough severe illnesses to overwhelm the medical care system sooner in many counties in the U.S.

The large variation in cases and deaths among counties raises the issue of the best strategies to reduce the spread of COVID-19 and other pathogens that are easily spread by human contact. A vigorous program of testing, tracing and quarantine, required masks and physical distancing in public places or resort to shut down in counties that are likely to have higher transmission rates as indicated by the data in this study may be more effective, efficient and less economically damaging than statewide shutdowns.

## Data Availability

I will put the data on my website, www.nanlee.net.

https:www.nanlee.net/countydata.xls

